# Reprogramming of the Sepsis N-Glycoproteome Illuminates a Functional Dissociation between Protein Abundance and Glycosylation in Immunothrombosis

**DOI:** 10.64898/2026.02.09.26345940

**Authors:** Dexiu Chen, Qidong Jiang, Zhangjing Shi, Yuxiang Yang, Liu Li, Xianying Lei, Chunxiang Zhang

## Abstract

**Purpose:** Sepsis-associated immunothrombosis significantly contributes to high mortality, yet the role of N-glycosylation in this process remains poorly understood. This study aimed to comprehensively profile the plasma N-glycosylation landscape in sepsis and elucidate how its specific reprogramming in the complement and coagulation cascades influences immunothrombotic balance and patient outcomes.

**Methods:** We performed in-depth 4D-DIA proteomic and N-glycomic analyses on plasma from 43 sepsis patients and 9 healthy controls. Differential expression, weighted gene co-expression network analysis (WGCNA), and protein–glycosylation correlation analyses were used to characterize molecular features. Clinical relevance was assessed via correlation and survival analyses.

**Results:** Extensive N-glycosylation reprogramming was observed in sepsis plasma,with marked enrichment in complement and coagulation pathways(KEGG p=7.76×10⁻ ²¹).Pro-coagulant proteins(eg,vWF,fibrinogen)showed increased abundance together with enhanced site-specific glycosylation,potentially amplifying their activity.In contrast,key anticoagulant proteins(eg,SERPINC1)displayed unchanged glycosylation at critical sites despite abundance changes,which may impair function.Survival analysis revealed distinct prognostic values of glycoproteins and specific glycosylation sites.For instance,high vWF protein levels predicted mortality(HR=2.83),whereas elevated glycosylation at vWF N211 was associated with improved survival(HR=0.135),suggesting a negative regulatory role.These glycosylation markers correlated closely with disease severity and prognosis,representing potential early-warning biomarkers independent of current clinical coagulation indicators.

**Conclusion:** Our study demonstrates widespread reprogramming of the plasma proteome and N-glycome in sepsis.We propose that decoupling of protein function from abundance through N-glycosylation in the complement-coagulation network contributes to immunothrombotic imbalance.Specific N-glycosylation sites may serve as novel prognostic biomarkers,offering new perspectives for early risk stratification and glycosylation-targeted therapies in sepsis.

**Key Points:**

1. Sepsis plasma exhibits specific N-glycosylation reprogramming overwhelmingly focused on the complement and coagulation cascade.
2. A dominant “glycosylation-dominated co-upregulation” mode in procoagulant factors, coupled with a “silent” glycosylation state in key anticoagulants, drives prothrombotic imbalance.
3. Site-specific N-glycosylation levels provide prognostic information distinct from, and often superior to, their carrier protein abundance, offering novel early-risk biomarkers.

## Introduction

Sepsis is defined as life-threatening organ dysfunction caused by a dysregulated host response to infection^[1]^.As a major public health challenge, sepsis imposes a heavy disease burden worldwide. In high-income countries, sepsis mortality ranges from 15% to 25%, and septic shock mortality can be as high as 30% to 40%, with even higher rates in low-income countries^[2]^. During the course of sepsis, most patients develop varying degrees of coagulation abnormalities. This process involves the coordinated action of fibrin, platelets, neutrophils, and monocytes, collectively leading to the formation of immunothrombosis^[3]^.This early response, to some extent, helps to identify and limit pathogen dissemination and tissue invasion, characterized by its local and self-limiting nature, typically without affecting overall organ perfusion^[4]^.However, once this balance is disrupted, the uncontrolled thrombotic inflammatory response can spread systemically, rapidly progressing to sepsis-induced coagulopathy (SIC) and even evolving into disseminated intravascular coagulation (DIC)^[5]^, ultimately resulting in multiple organ failure and uncontrollable bleeding. It is reported that approximately 30% to 50% of patients may further develop DIC, significantly increasing the risk of death^[6]^.

The formation of immunothrombosis in sepsis is a multisystem interactive process triggered by pathogen invasion. Its core mechanism begins with endothelial cell dysfunction: activated endothelial cells highly express tissue factor, initiating the extrinsic coagulation pathway, while downregulating thrombomodulin, leading to the inactivation of natural anticoagulant mechanisms. Circulating neutrophils are massively recruited and release neutrophil extracellular traps (NETs), whose DNA scaffold provides a framework for thrombus formation, and whose surface histones and granule proteins directly activate platelets and enhance thrombin generation.

Furthermore, excessive activation of the complement system, particularly C5a, acts as a potent inflammatory amplifier, further driving leukocyte activation and tissue factor expression^[3]^. The aforementioned pathological processes closely align with the complement and coagulation cascades pathway. This pathway integrates the three major systems of complement, coagulation, and kinin-kallikrein, revealing a positive feedback loop in sepsis triggered by immune activation and mediated through common molecular platforms, ultimately leading to uncontrolled thrombosis in the microvasculature^[6]^. Despite the close association between coagulation dysfunction and poor prognosis in sepsis patients, there are currently no effective treatments targeting the process of immunothrombosis. Various anticoagulants and antiplatelet agents have been evaluated in clinical trials, but their efficacy and safety remain controversial^[3, 5, 6]^. Therefore, further exploration of the regulatory mechanisms of immunothrombosis in sepsis holds significant clinical importance. The existing theoretical framework primarily focuses on protein expression, activation, and consumption, largely overlooking the fine-tuned regulation of protein function—an area where post-translational modifications may play a critical role.

In recent years, the rapid advancement of high-throughput proteomic technologies has provided unprecedented opportunities for systematically investigating post-translational modifications of proteins involved in immunothrombosis. In this study, we conducted a comprehensive analysis of plasma samples from sepsis patients and healthy controls using 4D-DIA-based proteomics and N-glycoproteomics. Our results demonstrate extensive reprogramming of the proteome and N-glycome in the plasma of sepsis patients, with these changes being significantly enriched in the complement and coagulation cascades pathway. Notably, the N-glycosylation levels of several key proteins closely correlate with patient disease severity, suggesting that protein N-glycosylation may have an important regulatory function in sepsis-associated immunothrombosis, independent of protein abundance. This provides a novel perspective for a deeper understanding of its pathological mechanisms.

## Methods

### 1. Ethics Approval and Patient Enrollment

#### 1.1 Ethical Statement and Study Design

This prospective cohort study was conducted in accordance with the Declaration of Helsinki and was approved by the Ethics Committee of the Affiliated Hospital of Southwest Medical University (Approval No. KY2025-390). Written informed consent was obtained from all participants or their legal representatives.

#### 1.2 Participant Recruitment and Enrollment

From July to October 2025, consecutive adult patients admitted to the intensive care unit (ICU) were screened. Sepsis was diagnosed according to the Sepsis-3 criteria (confirmed or suspected infection plus an increase in the Sequential Organ Failure Assessment [SOFA] score ≥ 2 points). Exclusion Criteria: For sepsis patients, exclusion criteria were: (1) age < 18 years; (2) pre-existing end-stage organ failure (e.g., chronic kidney disease stage 5 requiring dialysis, severe chronic liver failure); (3) decision to withdraw active treatment within 24 hours of admission. During this period, 51 patients met the diagnostic criteria. Of these, 8 were excluded: 6 left against medical advice and 2 were aged < 18 years. Consequently, 43 sepsis patients were enrolled within 24 hours of ICU admission. In parallel, 15 volunteers were screened from the hospital’s health examination center. Six were excluded due to active infection (n = 2), uncontrolled diabetes (n = 3), or declining participation (n = 1). Nine eligible individuals were enrolled as healthy controls. A total of 52 participants (43 sepsis, 9 controls) were included in the cross-sectional proteomic and glycomic analyses. All 43 sepsis patients were prospectively followed for 90-day survival.

#### 1.3 Sample Collection and Processing

Peripheral blood was collected in EDTA tubes. Plasma was separated by centrifugation at 2,000 × g for 15 minutes at 4°C, aliquoted, and stored at −80°C until analysis.

#### 1.4 Clinical Data Collection

Demographic information, comorbidities, SOFA and Acute Physiology and Chronic Health Evaluation II (APACHE II) scores at enrollment, and key laboratory parameters were collected prospectively and are summarized in Table S1.

### 2. Sample Preparation for Proteomic and N-Glycoproteomic Analysis

#### 2.1 Protein Extraction and Digestion

Plasma samples were thawed on ice in the presence of phosphatase and protease inhibitors (including 1 mM PMSF). Total protein was extracted by centrifugation at 12,000 × g for 10 min at 4°C. The supernatant was collected, and protein concentration was determined using a BCA assay.

For digestion, equal amounts of protein from each sample were reduced with 5 mM dithiothreitol (DTT) at 55°C for 30 min and alkylated with 10 mM iodoacetamide (IAA) in the dark at room temperature for 15 min. Proteins were then precipitated overnight at −20°C using six volumes of pre-cooled acetone. The pellet was collected by centrifugation at 8,000 × g for 10 min at 4°C, resuspended, and digested with trypsin at a 1:50 (enzyme-to-protein, w/w) ratio at 37°C for 12 h. The reaction was quenched by acidification with phosphoric acid to pH ∼3. The resulting peptides were desalted using Sep-Pak C18 cartridges and dried under vacuum.

#### 2.2 N-Glycopeptide Enrichment

Dried peptides were resuspended in 200 μL of loading buffer (85% acetonitrile, 1% formic acid) and loaded onto equilibrated ZIC-HILIC tips. After loading, the tips were washed twice: first with 40 μL of washing buffer 1 (82% acetonitrile, 1% formic acid) and then with 40 μL of washing buffer 2 (80% acetonitrile, 1% formic acid), with centrifugation at 2,000 × g for 30 s each. Enriched N-glycopeptides were eluted twice with 20 μL of elution buffer (10% ammonium hydroxide) by centrifugation at 6,000 × g for 30 s. The eluates were combined and lyophilized.

#### 2.3 Deglycosylation

Lyophilized N-glycopeptides were resuspended in 50 mM NH₄HCO₃ (prepared in H₂¹⁸O) and digested with 6 μL of PNGase F (also prepared in H₂¹⁸O) at 37°C for 2 h to release glycans and incorporate an ¹⁸O-label at the deamidated asparagine site.

### 3. Liquid Chromatography-Tandem Mass Spectrometry (LC-MS/MS) Analysis

Proteomic analysis was performed by Shanghai Luming Biotechnology Co., Ltd. Peptides were separated using a nanoflow UHPLC system (Vanquish Neo, Thermo Fisher Scientific) equipped with an analytical column (PepMap RSLC C18, 2 μm, 100 Å, 75 μm × 25 cm). Mobile phase A was 0.1% formic acid in water, and mobile phase B was 0.1% formic acid in 80% acetonitrile. The chromatographic gradient was as follows: 6–28% B over 27.5 min, 28–80% B over 1.5 min, held at 80% B for 1 min, and re-equilibrated to 6% B, with a total run time of 30 min.

MS data were acquired on a timsTOF HT mass spectrometer (Bruker Daltonics) coupled with an electrospray ionization (ESI) source. Data-independent acquisition (DIA) mode was used. The capillary voltage was set to 1750 V. Full MS scans ranged from 100 to 1700 m/z. Ion mobility separation was performed with a 1/K0 range of 0.6 to 1.6 V·s/cm².

### 4. Database Search and Data Processing

Raw DIA data were processed using Spectronaut Pulsar (version 18.7, Biognosys) against the UniProt human reference proteome database (release 2024_02). Search parameters included: fixed modification of carbamidomethylation (C); variable modifications of oxidation (M), acetylation (protein N-term), and deamidation (N, with ¹⁸O-label considered for N-glycosylation sites); trypsin digestion with up to two missed cleavages; precursor and protein Q-value cutoffs of 0.01. Protein quantification was performed at the MS2 level.

For N-glycosylation site analysis, sites were retained if they were quantified in at least two samples and in ≥50% of samples in at least one group. Missing values were imputed using the half-minimum value method within the data matrix. Data were normalized by median centering and log₂-transformed for subsequent analyses.

### 5. Bioinformatics and Statistical Analysis

All bioinformatics and statistical analyses were performed in R (version 4.4.3).

#### 5.1 Differential Expression and Enrichment Analysis

Differential expression analysis of proteins and N-glycosylation sites was performed using the limma package (v3.60.2). Significance thresholds were set at |log₂(fold change)| > 1 and false discovery rate (FDR)-adjusted p < 0.05. Functional enrichment analysis (Gene Ontology [GO], Kyoto Encyclopedia of Genes and Genomes [KEGG], Reactome) was conducted using the clusterProfiler package (v4.12.2), with FDR < 0.05 considered significant.

#### 5.2 Weighted Gene Co-expression Network Analysis (WGCNA)

WGCNA was performed using the WGCNA package (v1.72-5). To ensure scale-free topology, soft-thresholding powers were selected (β = 12 for the proteome, β = 8 for the N-glycome). Modules were identified with a merge cut height of 0.25. The correlation between module eigengenes and sepsis status was calculated to identify disease-relevant modules (e.g., the turquoise module), which were subsequently subjected to functional enrichment analysis.

#### 5.3 Association Pattern Analysis between Proteins and Glycosylation Sites

Proteins and their corresponding N-glycosylation sites with significant changes (fold change ≥ 2 or ≤ 1/2, p < 0.05 in both layers) were integrated. Based on their directional changes, four distinct association patterns were defined. Pathway enrichment analysis was performed for the protein sets within each pattern.

#### 5.4 Clinical Correlation and Survival Analysis

A systematic three-tier strategy was employed to screen for clinically relevant biomarkers: 1) Pathway focus: candidates were restricted to the “Complement and coagulation cascades” pathway (hsa04610); 2) Functional relevance: key hub, regulator, or effector molecules within the pathway were prioritized; 3)

Data-driven selection: molecules whose N-glycosylation sites exhibited “hyper-upregulated” or “hyper-downregulated” patterns were selected. The final core markers included C3, C7, C9, C8B, CFI, FGA, FGB, FGG, PROS1, SERPINA5, SERPING1, VWF, and their specific N-glycosylation sites.

Correlations between marker abundance (protein or glycosylation site) and clinical indices (e.g., SOFA, APACHE II, fibrinogen, renal/hepatic function markers, lactate) were assessed using Pearson or Spearman correlation via the psych package (v2.4.1).

For survival analysis, patients were dichotomized into high- and low-expression groups based on the median value of each marker. Cox proportional hazards regression was used to calculate hazard ratios (HRs) with 95% confidence intervals (CIs). Kaplan-Meier curves were plotted, and differences were assessed using the log-rank test. Due to the limited sample size and all mortality events occurring within 60 days, a sensitivity analysis using logistic regression for 60-day mortality was also performed ((Table S2). Analyses were conducted using the survival and survminer packages.

To examine the association with sepsis-induced coagulopathy (SIC), patients were stratified into SIC-positive and SIC-negative groups according to ISTH criteria. Differences in marker levels between groups were compared using the Wilcoxon rank-sum test or Student’s t-test.

All visualizations (PCA plots, volcano plots, enrichment plots, heatmaps, forest plots, survival curves) were generated using ggplot2 (v3.5.1) and pheatmap (v1.0.12). A two-sided *p* < 0.05 was considered statistically significant, with appropriate adjustments for multiple testing where applicable. **Results**

### 1. Extensive Reprogramming of the Plasma Proteome and N-Glycome in Sepsis

We performed 4D-DIA-based proteomic and N-glycoproteomic analyses on plasma samples from 43 sepsis patients and 9 healthy controls (HC). Baseline characteristics are listed in Table S1, showing no significant differences in age, sex, BMI, or comorbidities between the groups. Principal component analysis (PCA) of the N-glycome data revealed a clear separation between the sepsis and HC groups (Fig. 2A). During preliminary analysis, samples S21 and S22 were identified as outliers. After confirming their clinical diagnosis and excluding technical errors, we retained them in subsequent analyses, noting that their inclusion did not affect the robustness of the overall conclusions.

**Figure 1.**
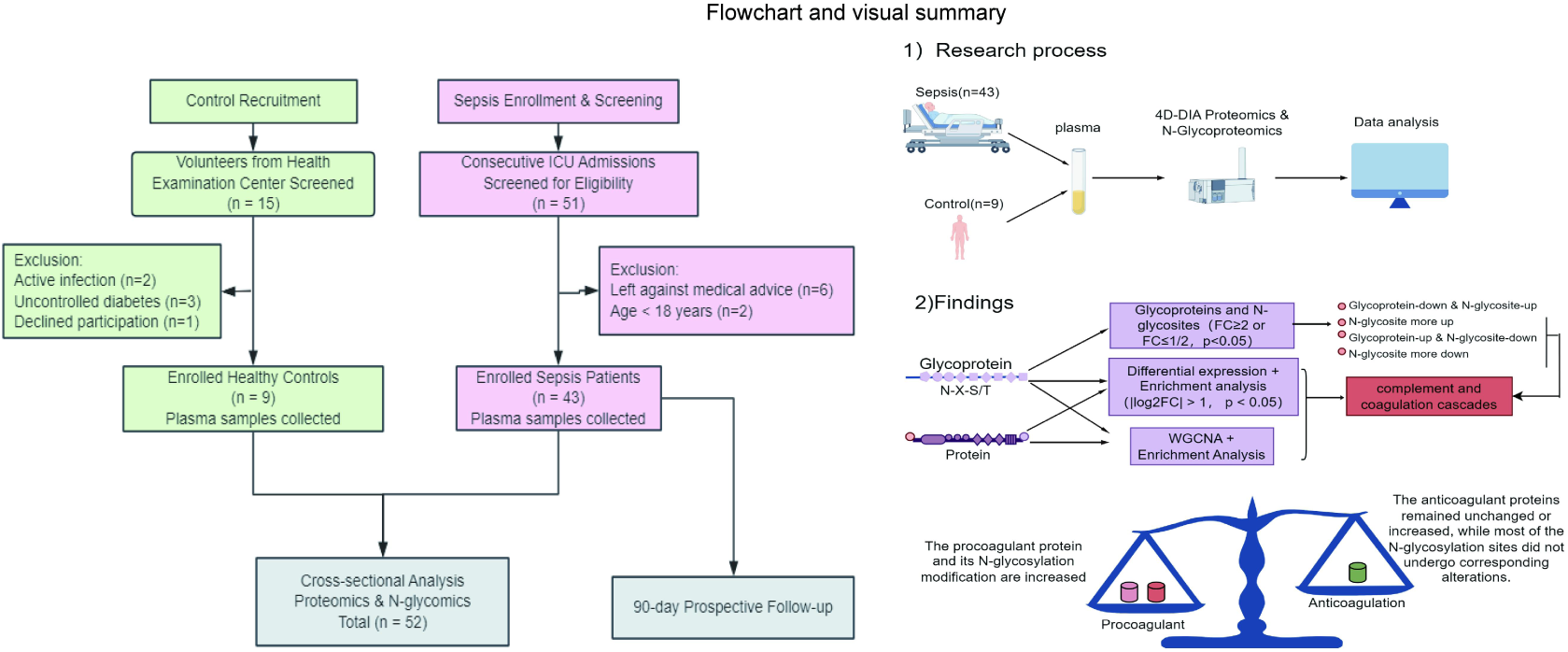
**Flowchart and visual summary**

**Figure 2.**
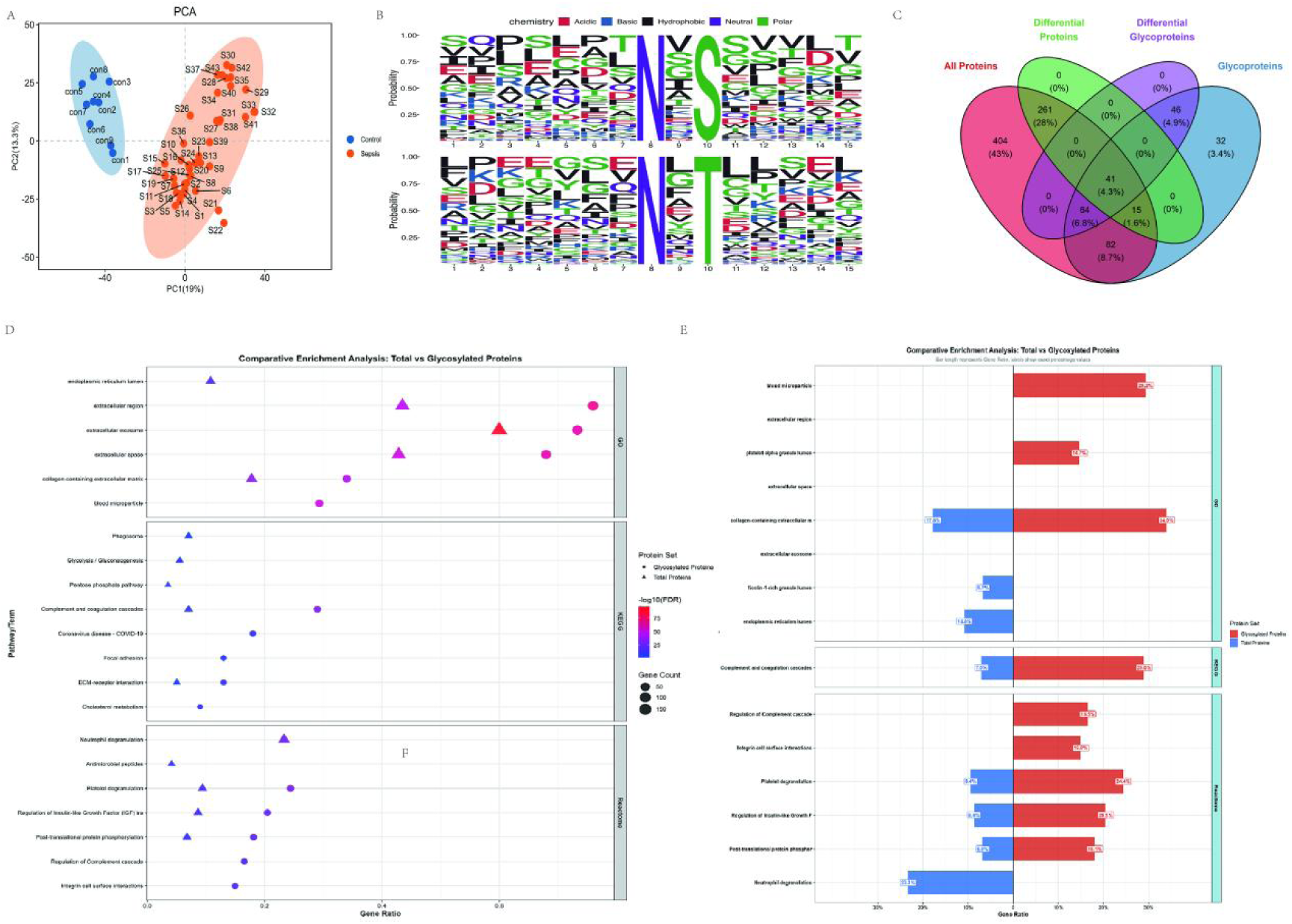
**Extensive Reprogramming of the Plasma Proteome and N-Glycome in Sepsis** (A) Principal Component Analysis (PCA) of the N-glycoproteome between the Healthy Control Group and the Sepsis Group. (B) Sequence characteristics of identified N-glycosylation sites. (C) A four-set Venn diagram illustrates the overlap among total proteins (n = 867), glycoproteins (n = 280), differentially expressed proteins (n = 317), and differentially expressed glycoproteins (n = 151). (D-E) .Comparison of enrichment analysis results of differentially expressed proteins and glycoproteins. (D) The position of the dots represents the gene ratio, the size corresponds to the number of enriched genes, the color represents the enrichment significance (−log10(FDR)), and the shape distinguishes the protein groups (triangle: protein; circle: glycoprotein). (E) Total proteins are shown on the left (blue), and glycoproteins are shown on the right (red). The length of the bars indicates the gene ratio, and the numerical labels show the exact percentage. The color saturation reflects the statistical significance (−log10(FDR)).

Motif analysis of the identified N-glycosylation sites confirmed strict adherence to the canonical N-X-S/T sequon (Fig. 2B), ensuring data quality^[7]^.

Notably, a stable enrichment of basic amino acid residues was observed in the upstream region (−7 to −3) of sepsis-associated glycosylation sites. The biological significance of this finding warrants further investigation but suggests that the sepsis microenvironment may be associated with a specific glycosylation modification preference.

A total of 867 proteins and 1172 specific N-glycosylation sites on 280 glycoproteins were quantified. Among these, 317 proteins and 151 glycoproteins (corresponding to 366 glycosylation sites) were differentially expressed between the sepsis and HC groups (|log₂FC| > 1, p < 0.05; Fig. 2C). Functional enrichment analysis showed that both differentially expressed proteins and glycoproteins were significantly enriched in pathways such as extracellular matrix organization and the complement and coagulation cascades. Crucially, glycoproteins demonstrated stronger pathway specificity (Fig. 2D). By calculating the ratio of enrichment scores between the N-glycome and proteome for specific pathways, we found that the enrichment significance of the N-glycome in the “Complement and coagulation cascades” pathway was approximately 314% higher than that of the proteome (Fig. 2E).

### 2. Functional Enrichment and Co-expression Network Analysis Jointly Identify the Complement/Coagulation Pathway as the Core Regulatory Axis of the N-Glycome

To systematically elucidate the expression patterns of proteins and glycoproteins in sepsis, we performed Weighted Gene Co-expression Network Analysis (WGCNA) on the proteomic and N-glycomic datasets separately.

Based on network topology properties, soft-thresholding powers of 12 (Fig. 3A) and 8 (Fig. 3B) were selected for constructing the proteomic and N-glycomic co-expression networks, respectively, achieving scale-free topology fit indices (R²) above 0.8. A dynamic tree cut height of 0.25 was set for module merging, and proteins or glycosylation sites with expression abundance standard deviation in the top 80% were retained for subsequent analysis.

**Figure 3.**
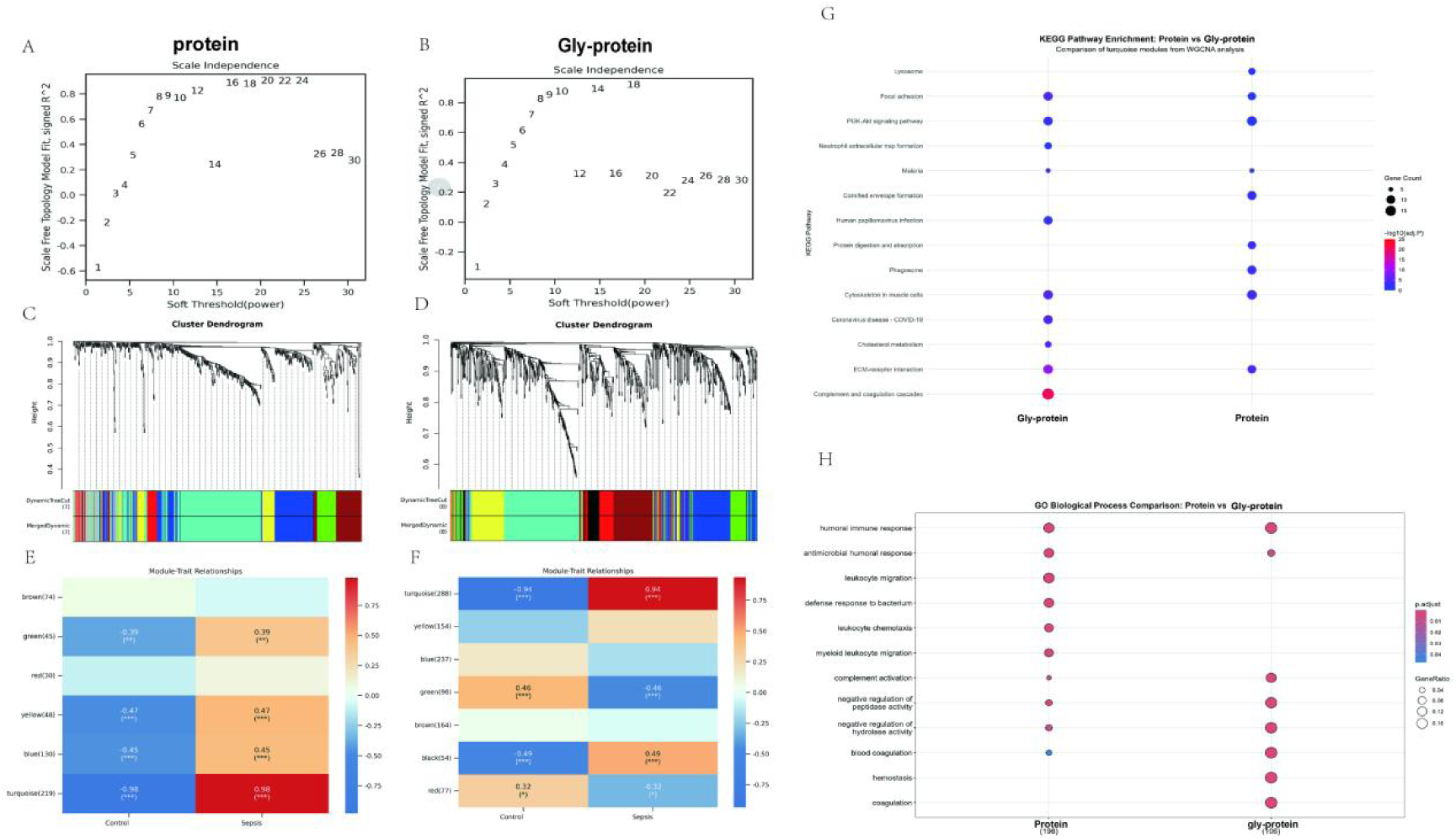
**By applying the WGCNA method, the distribution characteristics and enrichment features of proteins and glycoproteins were demonstrated.** (A, B) Represent the process of selecting the soft threshold power. The power for protein data selection is 12 (A), and for glycosylation protein data, it is 8 (B). Since at this value, the scale-free topological fitting index (R²) first reaches 0.8, indicating that the constructed network has good scale-free topological characteristics, which meets the analysis requirements of WGCNA. (C–F) The module-phenotype association analysis identified the core module significantly related to sepsis. In the protein group, the turquoise module (containing 237 proteins) has an extremely significant negative correlation with the sepsis state (r = −0.98) (C, E). In the glycoprotein group, the turquoise module (containing 288 glycosylation sites) also has an extremely significant negative correlation with the sepsis state (r = −0.94) (D, F). Comparative bubble plot of KEGG pathway enrichment. (H) Comparative bubble plot of significantly enriched Biological Process terms.(G,H)The size of the dot indicates the number of molecules in the term, and the color represents the adjusted p-value. *P < 0.05, **P < 0.01, ***P < 0.001.

Module-trait association analysis identified core modules highly correlated with sepsis status. In the proteomic network, the turquoise module (containing 237 proteins) showed a strong negative correlation with sepsis status (r = −0.98, p < 0.001). Similarly, in the N-glycomic network, the turquoise module (containing 288 glycosylation sites) also exhibited a strong negative correlation with sepsis status (r = −0.94, p < 0.001) (Fig. 3C–F). These two core modules were selected for in-depth functional analysis.

Functional analysis of the core modules showed that at the cellular component level, both were significantly enriched in locations such as the extracellular matrix, secretory granule lumen, and endoplasmic reticulum lumen (Fig. S1). Notably, the N-glycomic module was specifically enriched in components related to platelet alpha granules and extracellular vesicles, suggesting a potential specific role for N-glycosylation in platelet function and extracellular vesicle biology. KEGG pathway analysis further revealed a functional divergence between the two omics layers (Fig. 3G). The N-glycomic module showed an extremely significant enrichment in the “Complement and coagulation cascades” pathway (p = 7.76×10⁻ ²¹), indicating that protein N-glycosylation is a key regulatory layer for this pathway in sepsis. In contrast, the proteomic module was not significantly enriched in this pathway but was more focused on pathways related to cell adhesion, endocytosis, and tissue homeostasis, such as “ECM-receptor interaction” and “Phagosome”. Overlap in some pathways (e.g., “ECM-receptor interaction”) suggests their potential joint involvement in remodeling the tissue microenvironment during sepsis.

GO biological process enrichment results (Fig. 3H) further supported this functional specialization. The proteomic module’s functions were highly concentrated on the activation and execution of innate immune responses, with its most significantly enriched terms including “humoral immune response” (p = 3.51×10⁻ ¹⁵), “leukocyte migration” (p = 2.91×10⁻ ¹⁴), and “antibacterial humoral response” (p = 1.15×10⁻ ⁹), reflecting the complete process from immune signal activation to effector cell recruitment. Conversely, the N-glycomic module was specifically enriched in the regulation of coagulation and complement systems, with its most significant terms being “blood coagulation” (p = 1.38×10⁻ ¹⁶), “complement activation” (p = 4.89×10⁻ ¹⁶), and “fibrinolysis” (p = 3.94×10⁻ ¹⁴). Most critically, this module was also exceptionally significantly enriched in processes related to the “negative regulation of endopeptidase activity” (p = 8.14×10⁻ ¹⁶). This suggests that this glycosylation-associated co-expression network, while positively driving coagulation and complement activation, may also embed a crucial layer of negative feedback inhibition, thereby achieving complex and fine-tuned regulation during the pathophysiology of sepsis.

### 3. Diverse Association Patterns Reveal Dissociation and Coordination between Glycoprotein Abundance and N-Glycosylation Site Modification Levels

To deeply investigate the relationship between glycoprotein abundance and the relative abundance of their specific N-glycosylation sites in sepsis, we integrated analysis on molecular pairs that showed significant changes at both the glycoprotein and its corresponding glycosylation site levels (fold change ≥ 2 or ≤ 1/2, p-value < 0.05). Analysis of the corresponding volcano plot indicated that 76.4% of glycoprotein-site pairs remained relatively stable in sepsis. Among the pairs with significant changes, we identified four characteristic association patterns (Fig. 4A): The most prominent was the “Glycosylation-dominated coordinated upregulation” pattern (13.5%), followed by the “Dissociation” pattern featuring glycoprotein upregulation but glycosylation downregulation (5.5%). The “Glycosylation-dominated coordinated downregulation” pattern (3.5%) and the “Reverse” pattern featuring glycoprotein downregulation but glycosylation upregulation (1.2%) were relatively less common.

**Figure 4.**
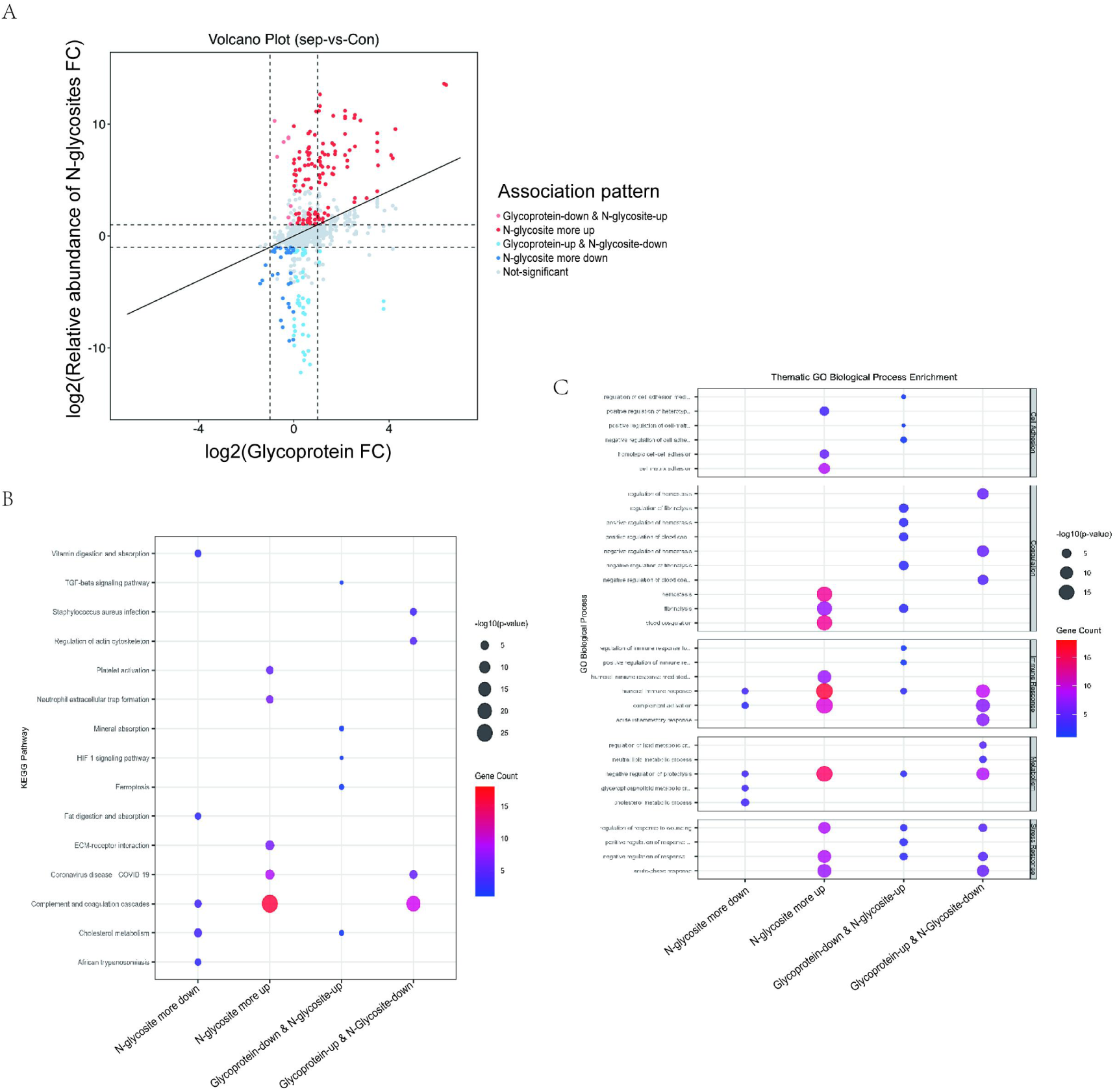
**The abundance of glycoproteins shows a diverse association pattern of decoupling and synergy with their N-glycosylation modifications.** A. Four association patterns of glycoprotein abundance and the occupancy of N-glycosylation sites in sepsis. (B, C) Enrichment analysis of glycoproteins for the four characteristic association patterns (B) and KEGG results (C).

Functional enrichment analysis of the glycoproteins within these four patterns revealed significant differences (Fig. 4B-C). Glycoproteins in the “Glycosylation-dominated coordinated upregulation” pattern were significantly enriched in pathways including the complement and coagulation cascades, neutrophil extracellular trap formation, and platelet activation. Glycoproteins in the “Glycosylation-dominated coordinated downregulation” pattern were primarily involved in lipid homeostasis-related processes such as lipoprotein particle remodeling, reverse cholesterol transport, phospholipid metabolism, and fat digestion and absorption. The “Dissociation” pattern (glycoprotein upregulation/glycosylation downregulation) was mainly associated with the negative regulation of the coagulation/fibrinolysis system, angiogenesis inhibition, and endothelial cell apoptosis regulation. The “Reverse” pattern (glycoprotein downregulation/glycosylation upregulation) was highly enriched in classical complement activation, acute inflammatory response, and B cell-mediated immunity. Overall, the complement and coagulation cascades pathway was significantly associated with glycoproteins across multiple patterns.

### 4. The Central Role and Specific N-Glycosylation Remodeling of the Complement and Coagulation Cascades

Building on the aforementioned findings—where differential expression analysis, WGCNA, and association pattern analysis all consistently pointed to the complement and coagulation cascades as the core pathological disturbance in sepsis—we conducted an in-depth mechanistic dissection of this pathway (Fig. 5). The results showed that the severe perturbation of this pathway was accompanied by specific and programmed changes in the N-glycosylation of key proteins.

**Figure 5.**
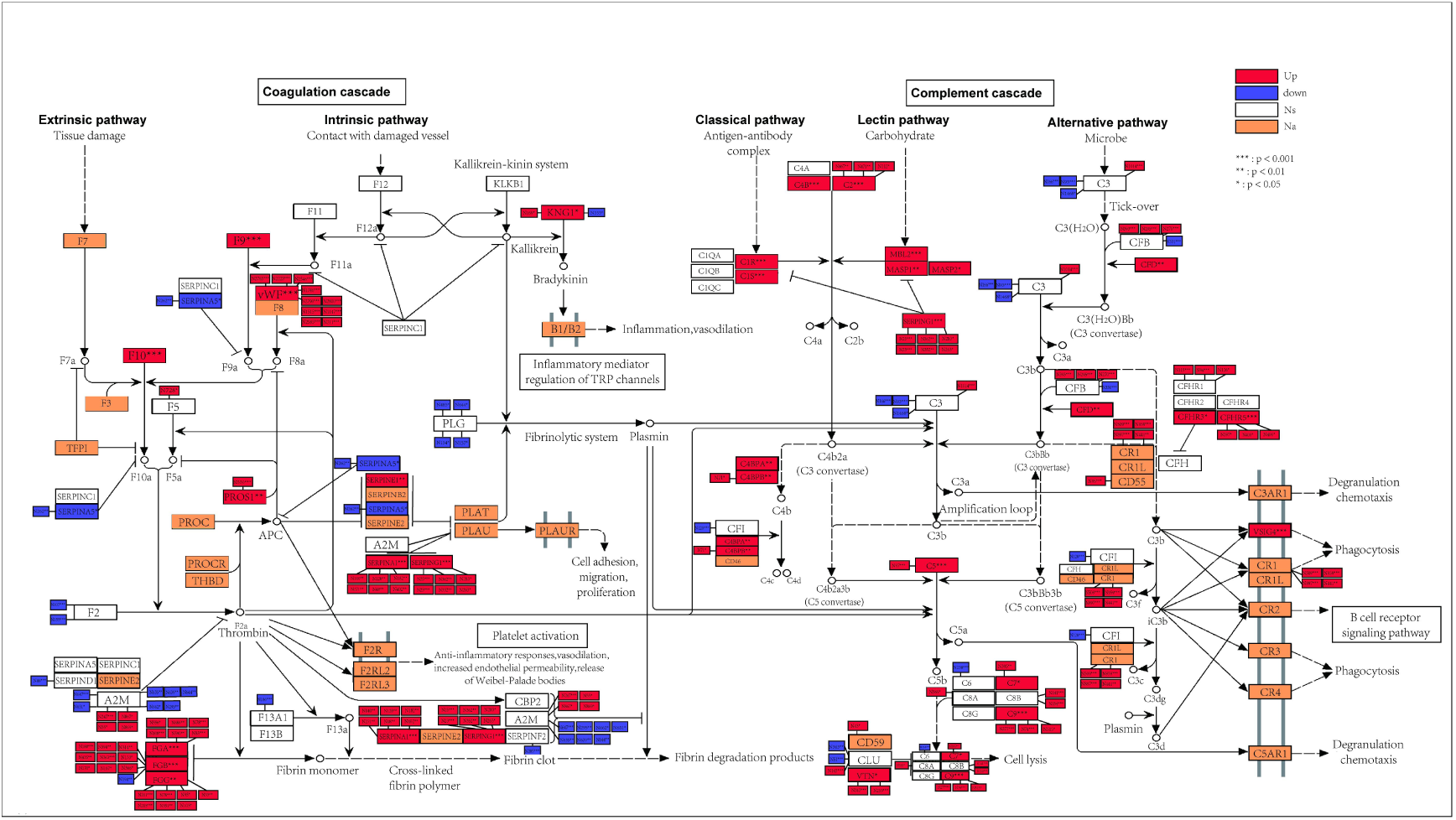
**Changes in proteins in the complement and coagulation cascade pathways and the specific N-glycosylation remodeling**

Within the coagulation system, we observed a functional enhancement of the procoagulant arm and a functional dissociation of the anticoagulant arm. The procoagulant arm exhibited a typical “N-glycosylation-dominated coordinated upregulation.” Protein levels of core factors like FIX, FX, vWF, and fibrinogen subunits (FGA, FGB, FGG) were generally elevated. Notably, for vWF and fibrinogen subunits, several key glycosylation sites (e.g., vWF N2790/N2800; FGB N363/N188; FGG N201/N215) showed an even more pronounced coordinated upregulation. This strongly suggests that glycosylation modifications may synergistically enhance the stability or functional activity of these procoagulant components, directly contributing to a hypercoagulable state. Concurrently, the anticoagulant mechanism displayed functional dissociation: while the protein C system cofactor PROS1 (both protein and its N530 site glycosylation) was upregulated, its key physiological inhibitor SERPINA5 showed a significant decrease in both protein and glycosylation levels (N262). Furthermore, the core anticoagulant protein SERPINC1 (antithrombin III) showed no change in its protein level or the glycosylation of all detected sites. This imbalanced regulatory pattern results in a relative insufficiency of anticoagulant reserve, unable to counterbalance the highly activated procoagulant process.

In the complement system, all three activation pathways were broadly activated, accompanied by specific N-glycosylation reprogramming. Multiple components of the classical (C1r, C1s), lectin (MBL2, MASP1/2), and terminal pathways (C5, C7, C8b, C9) showed upregulation at the protein level. Notably, the glycosylation of key terminal pathway components C7 (N395), C8B (N154, N141), and C9 (N277) was also significantly enhanced, suggesting that the assembly efficiency of the membrane attack complex (MAC) might be potentiated through glycosylation. Most crucially, the complement cascade amplification hub C3, while its total protein level remained unchanged, exhibited a significant upregulation in glycosylation at its N1014 site, fitting an “N-glycosylation-specific upregulation” pattern. This may potentially alter its cleavage kinetics or interactions with regulatory factors. In contrast, the glycosylation of major negative regulators CFI and CFH showed a decreasing or unchanged trend.

### 5. Correlation of Key Node Protein N-Glycosylation Levels in the Complement and Coagulation Cascades with Clinical Data

To screen for pathophysiologically significant markers, we employed a systematic three-tier strategy: 1) Focus on the complement and coagulation pathway consistent across enrichment analyses; 2) Prioritize functionally critical molecules within the pathway; and 3) Concentrate on sites exhibiting “hyper-upregulated” or “hyper-downregulated” N-glycosylation patterns. The screened core proteins and their corresponding sites included: C3, C7, C9, C8B, CFI, FGA, FGB, FGG, PROS1, SERPINA5, SERPING1, VWF, and their specific glycosylation sites.

The results showed that the glycosylation modifications of procoagulant components positively correlated with disease severity (Fig. 6). Both the protein levels of fibrinogen subunits (FGA, FGB, FGG) and their specific glycosylation sites (e.g., FGG_N215, FGG_N201) showed highly significant positive correlations with plasma fibrinogen concentration (e.g., FGG_N215: r = 0.521, p = 3.37×10⁻ ⁴), suggesting their “hyper-upregulation” may directly promote a hypercoagulable state. Similarly, vWF protein and several of its glycosylation sites (e.g., N1746, N2790, N2800) showed positive correlations with SOFA score, APACHE II score, and liver function markers (AST, ALT) (e.g., vWF protein vs. SOFA: r = 0.579, p = 4.82×10⁻ ⁵), highlighting the central role of glycosylated vWF in linking endothelial damage, coagulation disturbance, and end-organ dysfunction.

**Figure 6.**
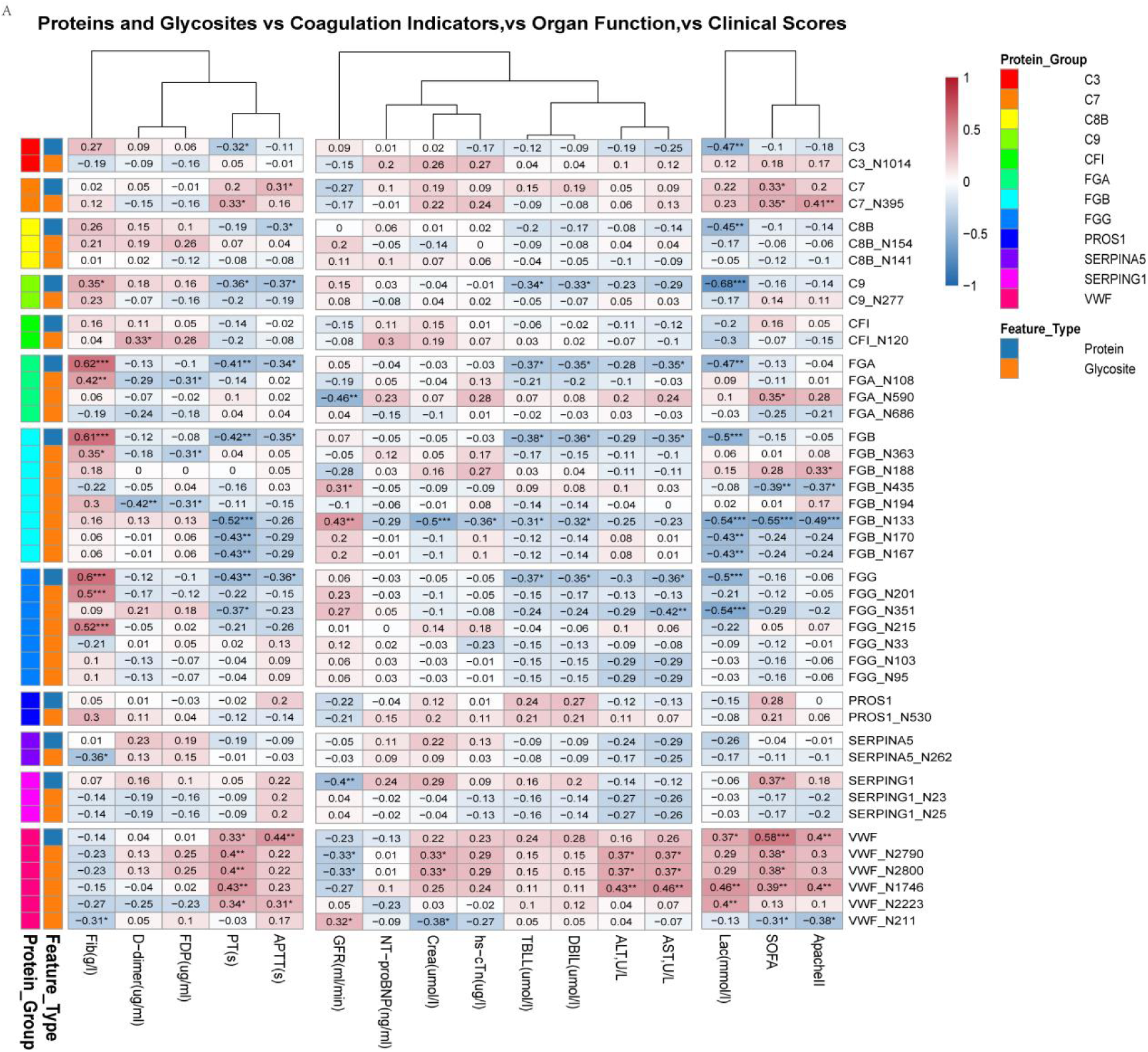
**Correlation of Key Node Protein N-Glycosylation Levels in the Complement and Coagulation Cascades with Clinical Data**

**Figure 6.**
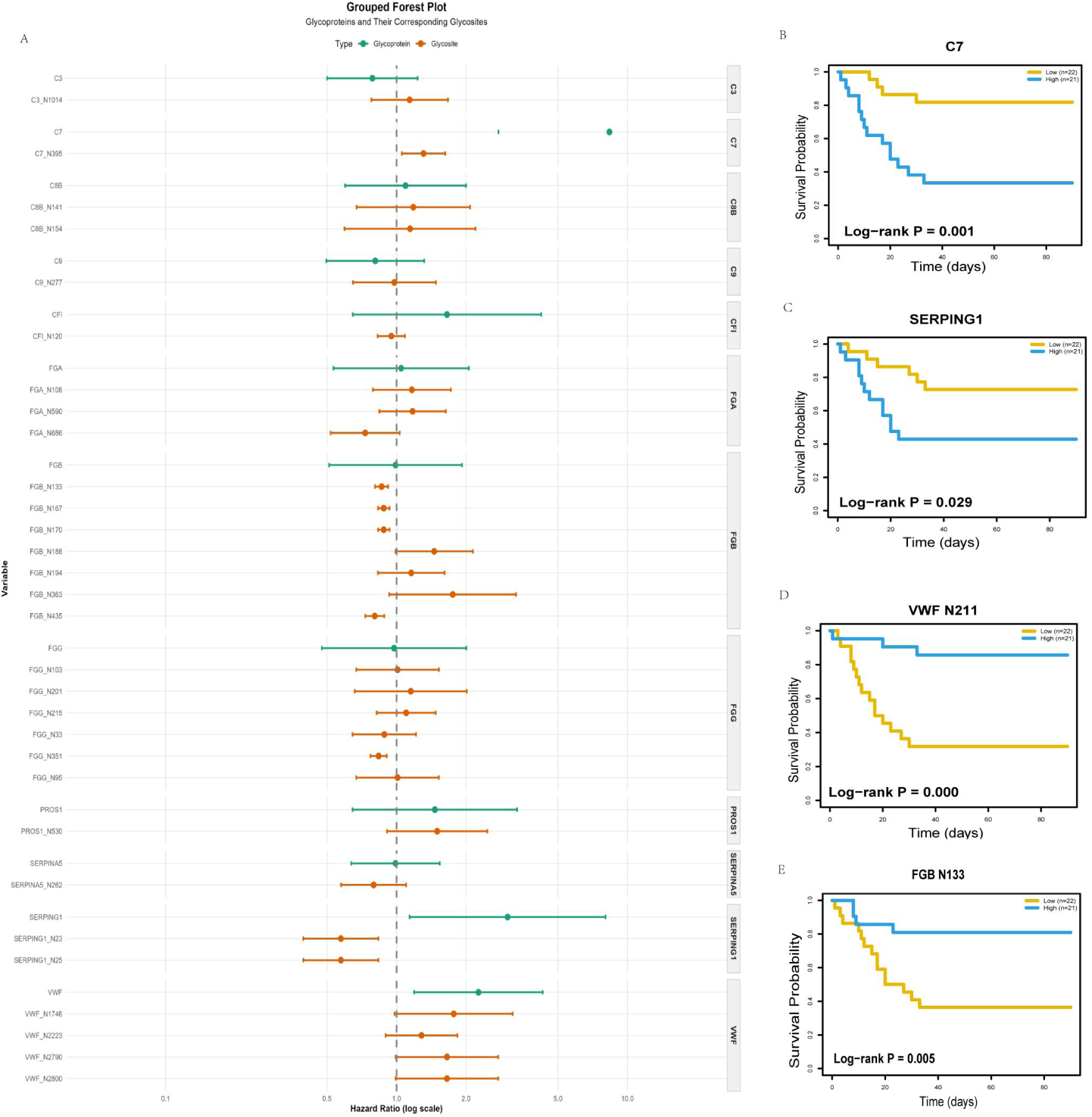
**Differential Prognostic Significance of Glycoproteins and Their N-Glycosylation Sites in Sepsis Survival** A.Forest plot of survival analysis of glycoproteins and their specific N-glycosylation sites.B-E.Kaplan-Meier survival curves for C7(B), SERPING1(C), VWF N211(D), and FGB N133(E).

Particularly noteworthy, we identified glycosylation sites associated with a protective effect. The N133 site on the FGB protein exhibited broad protective correlations: its glycosylation level was significantly negatively correlated with SOFA score, APACHE II score, blood lactate, creatinine, and prothrombin time (all p < 0.001). This suggests that this site may confer upon the fibrinogen beta chain a novel function related to improved organ perfusion and prognosis.

Conversely, the protein level of the terminal complement component C9 showed a strong negative correlation with blood lactate (r = −0.684, p = 4.36×10⁻ ⁷), possibly reflecting its excessive consumption in severe shock states. Furthermore, the glycosylation reprogramming of the complement system also correlated with disease severity: SERPING1 glycosylation negatively correlated with estimated glomerular filtration rate (eGFR) (r = −0.402, p = 0.00752), while C7_N395 glycosylation positively correlated with both APACHE II and SOFA scores (r > 0.406, p < 0.007). These findings collectively demonstrate that the specific N-glycosylation modifications of key nodes in the complement and coagulation pathways exhibit extensive and specific associations with the clinical manifestations and organ function injury in sepsis.

### 6. Differential Prognostic Value of Glycoproteins and Their Specific N-Glycosylation Sites for Sepsis Patient Survival

To systematically evaluate the prognostic value of core components within the complement and coagulation pathways, we performed survival analysis on the levels of 12 key glycoproteins and their 31 specific N-glycosylation sites. The analysis revealed that the abundance of the carrier protein and the site-specific glycosylation level provided prognostic information at different levels, sometimes even contradictory (Fig 6A). At the glycoprotein level, high abundance of C7 (HR = 5.49, 95% CI: 1.86–16.19, p = 0.002) (Fig 6B), PROS1 (HR = 2.78, 95% CI: 1.04–7.45, p = 0.042), SERPING1 (HR = 2.88, 95% CI: 1.07–7.72, p = 0.035) (Fig 6C), and VWF (HR = 2.83, 95% CI: 1.06–7.54, p = 0.038) was significantly associated with increased mortality risk.

Analysis of the relative abundance of the 31 specific N-glycosylation sites showed that 7 sites were significantly associated with 90-day prognosis in sepsis patients. Among them, high glycosylation abundance at the VWF_N211 site was a strong protective factor (HR = 0.135, 95% CI: 0.04–0.43, Cox P = 0.0017, Log-rank P = 0.0003) (Fig 6D). The 90-day mortality rate in the high-abundance group (14.3%) was significantly lower than that in the low-abundance group (68.2%). Increased glycosylation abundance at the

FGB_N133 (HR = 0.232, P = 0.0102) (Fig 6E) and SERPING1_N23/N25 (HR = 0.303, P = 0.0240) sites also showed protective effects. In contrast, increased glycosylation abundance at the C7_N395 (HR = 3.76, P = 0.0122) and FGB_N188 (HR = 3.85, P = 0.0108) sites were risk factors for patient prognosis.

### 7. Association Analysis of Core Glycosylation Markers with Sepsis-Induced Coagulopathy Phenotype

To further investigate whether these core glycosylation markers were specifically associated with the sepsis-induced coagulopathy (SIC) phenotype, we stratified the cohort into SIC-positive and SIC-negative groups based on the International Society on Thrombosis and Haemostasis (ISTH) criteria^[8]^.

The scoring system was as follows: Platelet count (≥150×10⁹/L: 0 points; 100–149×10⁹/L: 1 point; <100×10⁹/L: 2 points), Prothrombin time-international normalized ratio (PT-INR) (<1.2: 0 points; 1.2–1.4: 1 point; >1.4: 2 points),

Fibrinogen level (≥2.0 g/L: 0 points; <2.0 g/L: 1 point), and SOFA score (0: 0 points; 1: 1 point; ≥2: 2 points). A total score of ≥4 points was diagnostic for SIC. The results showed that despite the strong correlation of these markers with overall disease severity and mortality, their expression levels showed no statistically significant difference between the SIC-positive and SIC-negative groups (all adjusted p > 0.05, (Table S3).

## Discussion

Dysregulated coagulation, and particularly the uncontrolled thromboinflammatory response it drives, constitutes a core pathological process leading to organ failure and high mortality in sepsis^[3]^. While significant progress has been made in understanding the interplay between immune cells and endothelial cells^[3, 5]^, the molecular regulatory mechanisms, especially the precise role of post-translational modifications (PTMs) driving this process, remain incompletely defined. Protein N-glycosylation, a pivotal PTM, serves as a “molecular switch” that fine-tunes protein stability, activity, and interactions^[9]^. For the first time, this study employed integrated 4D-DIA plasma N-glycoproteomics and proteomics to systematically map the molecular landscape of sepsis, revealing a specific reprogramming of the complement and coagulation cascades. We propose a novel mechanism wherein the “functional dissociation of proteins from their abundance via N-glycosylation” drives the imbalance in immunothrombosis.

Our comprehensive analysis uncovered extensive molecular reprogramming in sepsis plasma. However, in contrast to the broad changes in the proteome, alterations in the N-glycome demonstrated remarkable functional focus. Both differential expression and co-expression network analyses converged, with extreme statistical significance, on the “complement and coagulation cascades” pathway. Crucially, the enrichment of this pathway at the N-glycosylation level far surpassed that observed for protein abundance changes (Fig. 2E, 3G). This clearly indicates that the functional regulation of this core pathological axis in sepsis occurs predominantly at the level of protein glycosylation, rather than through mere changes in expression quantity. This finding provides a direct molecular explanation, from a functional modification perspective, for previous transcriptomic studies linking early overactivation of coagulation and complement pathways to poor prognosis^[10]^. Delving deeper into the relationship between glycoproteins and their modification sites, we identified four distinct association patterns. Among these, the “N-glycosylation-dominated coordinated upregulation” pattern was predominant and specifically enriched in processes related to the complement/coagulation pathway, platelet activation, and neutrophil extracellular trap (NET) formation. This aligns perfectly with established sepsis pathophysiology: NETs serve as scaffolds for platelet activation and fibrin deposition^[11]^, and activated platelets release pro-thrombotic substances like vWF via alpha granules^[12]^. Our data show that key procoagulant components, such as vWF and fibrinogen subunits, not only exhibit elevated protein levels but also display more pronounced “hyper-upregulation” at multiple specific glycosylation sites (e.g., vWF N2790/N2800, FGG N201/N215). It is known that the glycosylation state of vWF directly influences its binding to platelet GP Ibα and its susceptibility to cleavage by ADAMTS13^[13, 14]^, while fibrinogen glycosylation can alter fibrin clot structure^[15]^. Therefore, we propose the “dual amplification” hypothesis: in sepsis, upregulated expression of procoagulant elements provides the substrate foundation, and the accompanying specific glycosylation “hyper-upregulation” may further amplify their pro-thrombotic activity by enhancing protein stability, optimizing interaction interfaces, or conferring protease resistance. A similar phenomenon has been reported, where the highly glycosylated form of CD147 is more effective in stimulating MMP expression and activity than its low-glycosylated counterpart^[16]^.

In sharp contrast to the “dual amplification” of the procoagulant system is the “functional dissociation and quiescence” of the anticoagulant system. The core physiological anticoagulant protein SERPINC1 (antithrombin III) showed no significant changes in its protein level or the glycosylation of all detected sites in sepsis. Given the crucial importance of N-glycosylation for its proper folding and anticoagulant activity^[17]^, this “quiescent” state implies that its functional reserve is not effectively mobilized. Concurrently, although the protein C system cofactor PROS1 was upregulated, its important inhibitor,

SERPINA5, showed a significant decrease in both protein and glycosylation levels. This asymmetric regulation—where the procoagulant arm is “glycosylation-amplified” while the anticoagulant arm remains in “functional quiescence”—constitutes a core molecular mechanism driving the precipitous tilt of the immunothrombotic balance toward a hypercoagulable state.

Our findings provide a new organizational framework for understanding the pathological “cross-talk” between the complement and coagulation systems. The coordinated upregulation of glycosylation on terminal complement pathway components (C7, C8B, C9) may enhance the assembly efficiency of the membrane attack complex. More importantly, while the total protein level of the complement cascade hub C3 remained unchanged, site-specific upregulation of glycosylation at its N1014 site could finely regulate its cleavage rate into C3a/C3b, thereby more efficiently driving downstream inflammatory and coagulation signals. Simultaneously, co-expression network analysis revealed that the glycosylation-related module, while positively driving coagulation and complement, was also significantly enriched in the “negative regulation of peptidase activity” (Fig. 3H). This indicates that the glycosylation reprogramming in sepsis is not a disorderly overactivation but rather a highly organized, adaptive (or maladaptive) process that encompasses both positive feedback amplification and an intrinsic intent for negative feedback counterbalance. Glycosylation, in this context, may serve as the key molecular language coordinating the synchronized activation of these two systems and attempting to set an activity “threshold.”

This series of molecular-level functional dysregulations ultimately manifests as novel biomarkers that are closely correlated with clinical severity and possess independent prognostic value. We found that levels of multiple glycosylation sites on procoagulant elements (e.g., vWF, fibrinogen) showed significant positive correlations with SOFA and APACHE II scores and markers of hepatic/renal injury, corroborating at the molecular modification level the pathological link between immunothrombosis and multi-organ dysfunction.

Particularly noteworthy is the identification of glycosylation events with potentially protective connotations: high glycosylation levels at the FGB_N133 site on the fibrinogen beta chain were associated with lower disease severity scores, better organ perfusion (lower lactate), and more normal coagulation function. This strongly suggests that different glycosylation sites on the same protein (FGB) (e.g., pro-thrombotic N188 vs. protective N133) may confer diametrically opposed biological effects through “site-specific competition” or conformational regulation.

Intriguingly, these glycosylation markers were independent of the SIC diagnosis, showing no difference between SIC-positive and SIC-negative groups, yet they were able to predict mortality risk across the entire sepsis cohort. Based on this, we posit that these glycosylation markers capture “molecular functional dysregulation” that precedes the appearance of abnormalities in conventional coagulation laboratory tests. They do not reflect established coagulopathy meeting diagnostic criteria but rather serve as an early warning of impending coagulation and endothelial dysfunction, thus possessing the unique advantage of being “sentinel” biomarkers. The most compelling example illustrating this “function-abundance dissociation” paradigm is VWF: a high level of VWF protein is a clear risk factor for death, whereas high glycosylation at its N211 site is strongly associated with improved survival. This suggests that glycosylation at this site may negatively regulate the pro-thrombotic activity of VWF. Compared to total protein levels, these site-specific modifications provide differentiated and potentially more precise prognostic information, a value already established in fields like oncology, where specific glycoforms are often superior biomarkers to their carrier proteins^[18]^.

This study systematically delineates the specific N-glycosylation reprogramming of the complement and coagulation cascades in sepsis plasma, moving beyond the traditional framework of protein abundance analysis. We propose for the first time that the “functional dissociation of proteins from their abundance via glycosylation” is a core mechanism of immunothrombosis in sepsis, specifically manifested as the asymmetric pattern of “glycosylation dual amplification” in the procoagulant system and “functional quiescence” in the anticoagulant system. The specific N-glycosylation sites identified in this study, serving as early molecular warning markers independent of current clinical coagulation diagnostics, open a new perspective for sepsis risk stratification.

Furthermore, the identification of key sites (e.g., VWF_N211) provides unprecedented candidate targets for developing precise antithrombotic therapies targeting glycosylation regulation.

We acknowledge several limitations of this study. First, this is primarily a discovery and association study; the precise molecular mechanisms underlying the function of key identified glycosylation sites (e.g., FGB_N133) await further validation through functional experiments (e.g., site-specific mutagenesis, glycosylation-engineered proteins). Second, being a single-center cohort with a limited sample size, these promising glycosylation prognostic markers require independent validation in larger-scale, multi-center prospective cohorts. Additionally, our results provide relative quantification of proteins and their N-glycosylation sites but do not definitively establish the site-specific glycosylation occupancy or elucidate the detailed glycan structure and composition.

In conclusion, our findings systematically reveal, for the first time, that protein N-glycosylation reprogramming is a core, yet long-overlooked, driver of immunothrombotic imbalance in sepsis. These discoveries not only provide a new “functional regulation” perspective that transcends protein abundance for understanding sepsis pathology but, more importantly, lay a solid foundation for developing novel prognostic tools and precision treatment strategies based on glycosylation modifications.

## Data Availability

The mass spectrometry proteomics and glycoproteomics raw data generated in this study have been deposited in the OMIX repository at China National Center for Bioinformation under accession number OMIX013679. The data are currently under private access for peer review. Reviewers can access the data via the following secure, time-limited link: https://ngdc.cncb.ac.cn/omix/preview/oZxQYPdN. The data will be made publicly available upon publication of this article.

## Acknowledgments

This work was supported by National Natural Science Foundation of China (Grant Nos. U23A20398; Grant Nos. 82030007), Noncommunicable Chronic Diseases-National Science and Technology Major Project (Grant Nos. 2024ZD0537707), Sichuan Science and Technology Program (Grant No. 2022YFS0632) and the Strategic Cooperation Project between Luzhou Municipal People’s Government and Southwest Medical University (Grant No. 2024LZXNYDR010).

We thank the Shanghai Luming biological technology co., LTD (Shanghai, China) for providing N-glycoproteomics/proteomics services.Conflicts of Interest All authors have completed the ICMJE uniform disclosure form. All authors declare that they have no financial or personal relationships with other people or organizations that could inappropriately influence (bias) their work. No potential conflicts of interest exist with respect to the research, authorship, and/or publication of this article.

